# Clinical, laboratory, and temporal predictors of neutralizing antibodies to SARS-CoV-2 after COVID-19

**DOI:** 10.1101/2020.10.06.20207472

**Authors:** Jim Boonyaratanakornkit, Chihiro Morishima, Stacy Selke, Danniel Zamora, Sarah McGuffin, Adrienne E. Shapiro, Victoria L. Campbell, Christopher L. McClurkan, Lichen Jing, Robin Gross, Janie Liang, Elena Postnikova, Steven Mazur, Anu Chaudhary, Marie K. Das, Susan L. Fink, Andrew Bryan, Alex L. Greninger, Keith R. Jerome, Michael R. Holbrook, Terry B. Gernsheimer, Mark H. Wener, Anna Wald, David M. Koelle

## Abstract

**Background:** SARS-CoV-2-specific antibodies may protect from reinfection and disease, providing the rationale for administration of plasma containing SARS-CoV-2 neutralizing antibodies (nAb) as a treatment for COVID-19. The clinical factors and laboratory assays to streamline plasma donor selection, and the durability of nAb responses, are incompletely understood.

**Methods:** Adults with virologically-documented SARS-CoV-2 infection in a convalescent plasma donor screening program were tested for serum IgG to SARS-CoV-2 spike protein S1 domain, nucleoprotein (NP), and for nAb.

**Results:** Amongst 250 consecutive persons studied a median of 67 days since symptom onset, 243/250 (97%) were seropositive on one or more assays. Sixty percent of donors had nAb titers ≥1:80. Correlates of higher nAb titer included older age (adjusted OR [AOR] 1.03/year of age, 95% CI 1.00-1.06), male sex (AOR 2.08, 95% CI 1.13-3.82), fever during acute illness (AOR 2.73, 95% CI 1.25-5.97), and disease severity represented by hospitalization (AOR 6.59, 95% CI 1.32-32.96). Receiver operating characteristic (ROC) analyses of anti-S1 and anti-NP antibody results yielded cutoffs that corresponded well with nAb titers, with the anti-S1 assay being slightly more predictive. NAb titers declined in 37 of 41 paired specimens collected a median of 98 days (range, 77-120) apart (*P*<0.001). Seven individuals (2.8%) were persistently seronegative and lacked T cell responses.

**Conclusions:** Nab titers correlated with COVID-19 severity, age, and sex. Standard commercially available SARS-CoV-2 IgG results can serve as useful surrogates for nAb testing. Functional nAb levels were found to decline and a small proportion of COVID-19 survivors lack adaptive immune responses.

## INTRODUCTION

Since its emergence in late 2019, Severe Acute Respiratory Syndrome Coronavirus 2 (SARS-CoV-2) infection has spread globally, with devastating effects in many communities. At this time, no vaccines or therapeutics for the prevention or treatment of SARS-CoV-2 infection have been approved by the Food and Drug Administration (FDA). Passive immunotherapy with convalescent plasma, hyperimmune gamma globulin, or monoclonal antibodies is beneficial for treatment or prophylaxis of several infections(1), and these approaches are under investigation in patients with COVID-19. Both single donor and pooled immunoglobulin products currently prioritize collection of convalescent donor plasma with high levels of neutralizing antibodies (nAb).

NAb assays are challenging to perform and have biosafety concerns. A better understanding of the predictors and correlates of high nAb titers could be useful for improving access to high quality therapeutic plasma products. In addition, it is uncertain how durable acquired immunity will be in response to SARS-CoV-2 infection. Protection from several other respiratory viruses after natural infection is temporary and incomplete(2, 3) and longitudinal information concerning nAb levels may assist modeling the future of the outbreak until effective vaccines become widely used. We evaluated antibody levels in candidate plasma donors recovered from virologically-documented SARS-CoV-2 infection and examined the trajectory of antibody levels in a subset of donors over time. Further, we compared nAb titers with two rapid commercially available assays to determine whether these assays could substitute for time- and labor-intensive nAb measurements that require a BSL-3 facility.

## RESULTS

### Participant characteristics

We enrolled 250 consecutive persons interested in convalescent plasma donation with a median age of 51 years (range 19-91); 48% were men (**Table 1**). The median duration of COVID-19 symptoms was 15 days (range 0-49), and the median number of days between positive PCR testing and initial antibody testing was 61 days (range 14-112). Eighteen (11%) of participants reported hospitalization including 9 (4%) who received ICU-level care. The most common symptoms reported were fatigue (86%), fever (74%), muscle aches (73%), cough (72%), anosmia/ageusia (61%), and headache (60%) (**Supplemental Figure 1A**). The most commonly reported underlying comorbidities were lung disease (14.4%), heart disease (13.6%), hypertension (12.8%), cancer (12.8%), hyperlipidemia (9.6%), and diabetes (9.6%) (**Supplemental Figure 1B**).

**Table 1.** Demographic and clinical characteristics of study subjects.

| Category | Value (N=250) |
| --- | --- |
| Age (range) | 51 (19-91) |
| Men | 119 (47.6%) |
| Hispanic ethnicity | 11 (4.4%) |
| Race |  |
| White | 214 (85.6) |
| Asian | 22 (8.8%) |
| Black | 4 (1.6%) |
| Pacific Islander | 1 (0.4%) |
| Native American | 1 (0.4%) |
| Other | 4 (1.6%) |
| More than one race | 4 (1.6%) |
| Median (range) days between symptom resolution and antibody test<sup>A</sup> | 50 (21-110) |
| Median (range) days between positive PCR and antibody test | 61 (14-112) |
| Severity |  |
| Outpatient<sup>B</sup> | 223 (89.2%) |
| Hospitalized but not in ICU | 18 (7.2%) |
| ICU | 9 (3.6%) |

<sup>A</sup>Excluding one asymptomatic person
<sup>B</sup>Includes one asymptomatic participant who was a household contact of a person with symptomatic COVID-19

### Clinical correlates of neutralizing antibody response

Among the 250 participants, 12.8% had nAb titers below 1:40, 27.6% had titers of 1:40, 34.4% had titers of 1:80, and 25.2% had titers of 1:160 or greater (**Figure 1)**. Next, we examined the clinical correlates of nAb titers of 1:80 or greater, as this group was eligible for donation to a pooled immunoglobulin product protocol. In a univariable analysis, older age (OR 1.04 per year, 95% CI 1.02-1.06), male sex (OR 2.27, 95% CI 1.35-3.82), the presence of fever (OR 3.12, 95% CI 1.73-5.61) or difficulty breathing (OR 2.12, 95% CI 1.26-3.54) during the acute illness episode, hospitalization denoting more severe disease (OR 9.98, 95% CI 2.31-43.16), and having diabetes (OR 5.36, 95% CI 1.55-18.48) were significantly associated with higher nAb titers (**Table 2**). In a multivariable analysis, older age (adjusted OR [AOR] 1.03 [95% CI 1.00-1.06]), male sex (AOR 2.08 [95% CI 1.13-3.82]), the presence of fever during the acute illness episode (AOR 2.73 [95% CI 1.25-5.97]), and hospitalization (AOR 6.59 [CI 1.32-32.96]) remained significantly associated with higher nAb titers (**Table 2**). A longer interval in time between the diagnostic PCR and antibody testing was also associated with lower nAb titers (OR 0.97 [CI 0.96-0.99]) **(Table 2)**.

**Figure 1.**
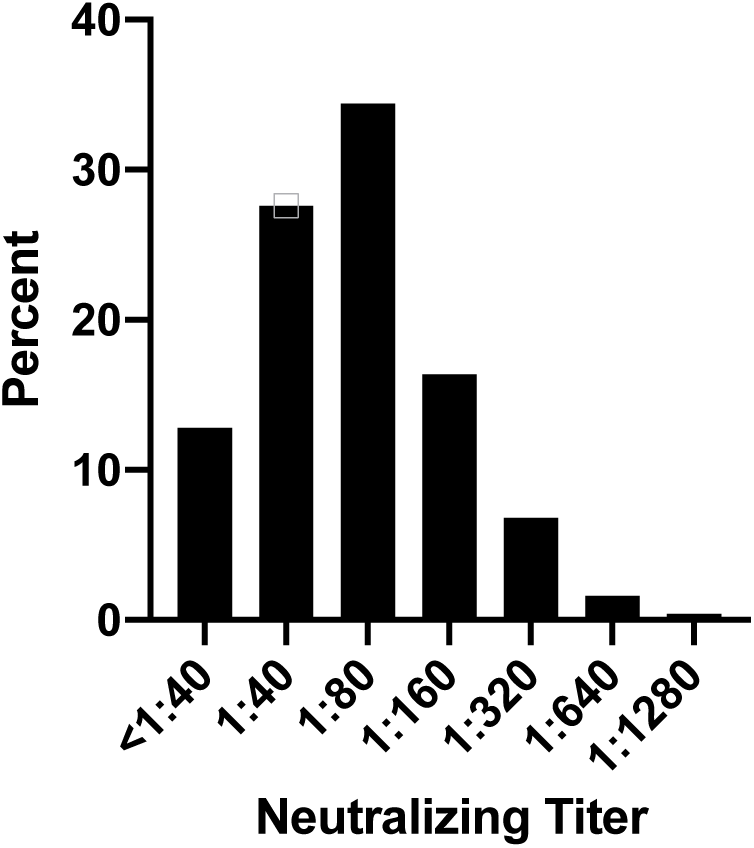
Distribution of neutralization antibody titers in convalescent subjects (N=250). Values on the x-axis represent the dilution factor of serum that yielded a 50% reduction in SARS-CoV-2 infection of Vero E6 cells.

**Table 2.** Clinical correlates of high (≥1:80) circulating SARS-CoV-2 neutralizing antibody titers.

| Co-variates | Univariable |  | Multivariable |  |
| --- | --- | --- | --- | --- |
|  | Odds Ratio (OR) | 95% Confidence Interval (CI) | Adjusted Odds Ratio (AOR) | 95% Confidence Interval (CI) |
| Age, per year | **1.04** | 1.02-1.06 | **1.03** | 1.00-1.06 |
| Men vs women | **2.27** | 1.35-3.82 | **2.08** | 1.13-3.82 |
| Race, white vs non-white | 0.52 | 0.24-1.13 |  |  |
| Symptoms |  |  |  |  |
| Fever | **3.12** | 1.73-5.61 | **2.73** | 1.25-5.97 |
| Cough | **1.52** | 0.87-2.67 | 0.94 | 0.45-1.97 |
| Runny nose | 0.97 | 0.57-1.65 |  |  |
| Difficulty breathing | **2.12** | 1.26-3.54 | 1.65 | 0.84-3.25 |
| Wheezing | 1.34 | 0.71-2.55 |  |  |
| Sore throat | **0.63** | 0.37-1.07 | 0.80 | 0.41-1.58 |
| Diarrhea | 0.83 | 0.49-1.40 |  |  |
| Fatigue | 0.86 | 0.41-1.80 |  |  |
| Muscle aches | 1.23 | 0.69-2.17 |  |  |
| Anosmia/ageusia | 0.77 | 0.45-1.30 |  |  |
| Headache | 0.99 | 0.59-1.67 |  |  |
| Number of symptoms | 1.07 | 0.95-1.21 | 1.08 | 0.86-1.34 |
| Days between positive PCR and antibody test | **0.98** | 0.97-0.99 | **0.97** | 0.96-0.99 |
| Duration of symptoms (days)<sup>A</sup> | 1.00 | 0.98-1.03 |  |  |
| Hospitalization | **9.98** | 2.31-43.16 | **6.59** | 1.32-32.96 |
| Comorbidities |  |  |  |  |
| Cancer | **2.23** | 0.96-5.19 | 2.27 | 0.81-6.38 |
| Heart disease | 1.49 | 0.69-3.22 |  |  |
| Kidney disease | 0.78 | 0.25-2.39 |  |  |
| Hypertension | 1.34 | 0.62-2.92 |  |  |
| Hyperlipidemia | 1.73 | 0.69-4.34 |  |  |
| Lung disease | 0.72 | 0.36-1.47 |  |  |
| Diabetes | **5.36** | 1.55-18.48 | 2.75 | 0.69-10.95 |

<sup>A</sup>excluding one asymptomatic subject
**Bolded** odds ratios indicate $P < 0.2$ in the univariable analysis or $P < 0.05$ in the multivariable analysis.

### Comparison of anti-SARS-CoV-2 antibody measurements

Circulating IgG antibodies against SARS-CoV-2 antigens were measured using IgG immunoassays for anti-S1 (Euroimmun) and anti-NP (Abbott). Amongst the 250 sera analyzed at the initial time point, 23 (9.2%) of samples were negative by the manufacturer’s criteria versus 11 (4.4%) by a z-score threshold of 3.0 for the Abbott assay. For the Euroimmun assay, 20 (8.0%) were negative by a z-score threshold of 3.0 versus 30 (12.0%) by the manufacturer’s criteria. A minority of samples demonstrated discordant results (**Figure 2**); overall, 15 (6%) were discordant for seropositivity using z-score thresholds of 3.0 versus 19 (7.6%) using manufacturers’ thresholds.

**Figure 2.**
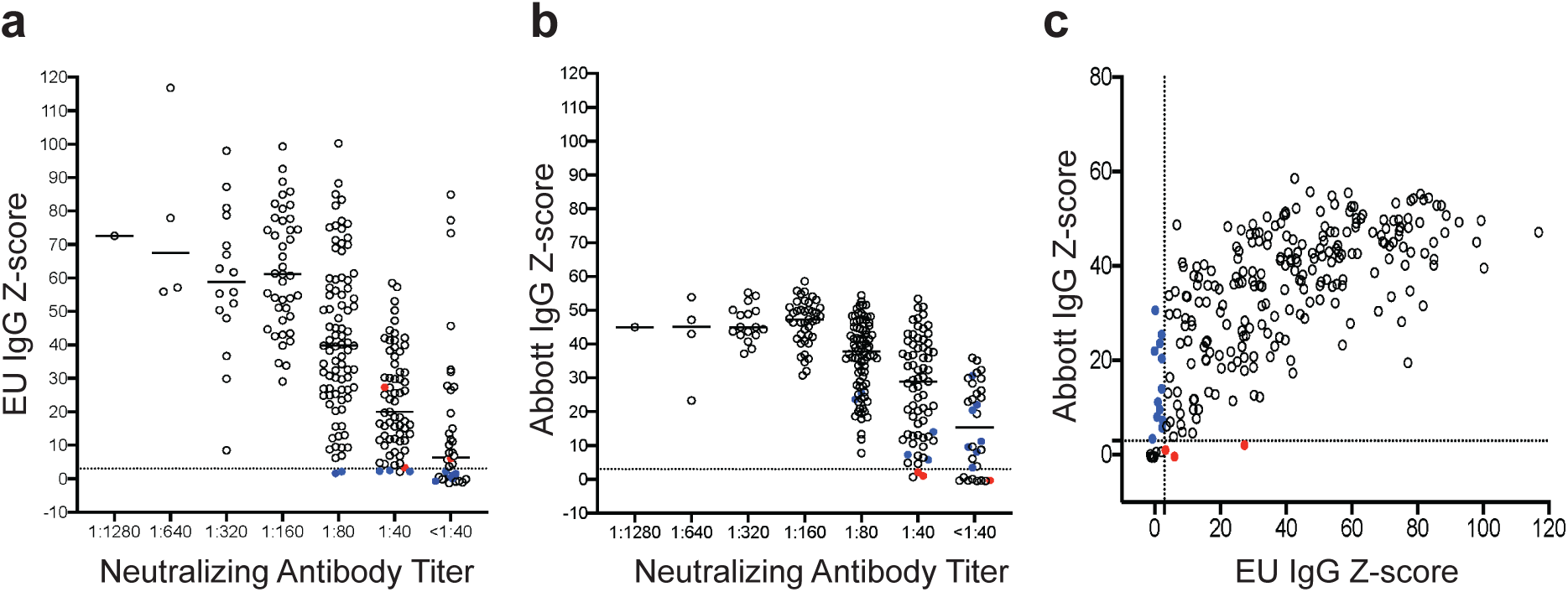
Relationship between serum measurements of anti-SARS-CoV-2 antibodies. Levels of anti-SARS-CoV-2 IgG antibodies specific for S1(Euroimmun, EU) (A) or of anti-SARS-CoV-2 IgG antibodies specific for NP (Abbott) expressed as z-scores (B) are shown relative to neutralizing antibody titers obtained from the same samples. Each symbol represents the initial specimen from one person. (C) Anti-SARS-CoV-2 S1 IgG levels (EU) relative to anti-SARS-CoV-2 nucleocapsid (N) protein IgG levels (Abbott). The dotted lines indicate the positivity threshold (z-score of 3) for both EU and Abbott assay results. Colored circles indicate discordant results between the Euroimmun and Abbott assays. Blue circles represent samples with negative z-score results (z-score <3.0) by Euroimmun and positive (z-score ≥3.0) by Abbott. Conversely, red circles represent samples with positive (z-score ≥3.0) results by Euroimmun and negative (z-score <3.0) by Abbott.

Quantitative assay results were generally concordant. Neutralizing antibody titers correlated with z-scores from the anti-S1 Euroimmun assay (Pearson correlation of 0.49, *P*<0.001) and with the Abbott anti-NP assay (Pearson correlation of 0.39, *P*<0.001) (**Figure 2A, B**). Z-scores of the Euroimmun and Abbott assays also correlated with each other (Pearson correlation of 0.69, *P*<0.001) (**Figure 2C**). Receiver operating characteristic (ROC) curves were generated to summarize the sensitivity and specificity of the Abbott and the Euroimmun assays for detecting nAb titers ≥ 1:40, ≥ 1:80 and ≥1:160 (**Supplemental Figure 2**). The area under the curve (AUC) for the Euroimmun assay was slightly greater than for the Abbott assay across all three nAb levels.

The criteria for positivity in the binding IgG tests was subjected to further analysis focusing on nAb cutoffs of ≥ 1:80 or higher. A z-score of 3.0 in the Euroimmun assay had a sensitivity of 99% and specificity of 17% for a nAb titer ≥ 1:80 (**Table 3 and Supplemental Figure 2B**). To estimate optimal cutoffs, we used Youden’s index(4), which indicated that at an optimized z-score cut-off of 34.2, the Euroimmun assay had a sensitivity of 78% and specificity of 72%. A z-score of 3.0 in the Abbott assay had a sensitivity of 100% but a specificity of only 11%. Using an optimized z-score cut-off of 32.6, the Abbott assay had a sensitivity of 77% and specificity of 81%. Because the optimal therapeutic titer of convalescent plasma is unknown, we also calculated the sensitivity and specificity of both IgG binding assays to predict nAb titers ≥ 1:160; as expected, these z-scores were higher (**Table 3**).

**Table 3.** Sensitivity and specificity of circulating IgG anti-S1 and anti-NP immunoassays for predicting neutralizing antibody titers.

| | nAb ≥ 1:80 | | | | nAb ≥ 1:160 | | | |
| --- | --- | --- | --- | --- | --- | --- | --- | --- |
| Criteria | Z-score | ODR | Sensitivity | Specificity | Z-score | ODR | Sensitivity | Specificity |
| Manufacturer's cut-off<sup>A</sup> |  |  |  |  |  |  |  |  |
| Anti-S1 IgG | 4.2 | 0.8 | 0.99 | 0.24 | 4.6 | 0.8 | 1.00 | 0.14 |
| Anti-NP IgG | 9.4 | 1.4 | 0.99 | 0.21 | 8.4 | 1.4 | 1.00 | 0.12 |
| Z-score cut-off of 3.0 |  |  |  |  |  |  |  |  |
| Anti-S1 IgG | 3.0 | 0.7 | 0.99 | 0.17 | 3.0 | 0.6 | 1.00 | 0.11 |
| Anti-NP IgG | 3.0 | 0.5 | 1.00 | 0.11 | 3.0 | 0.5 | 1.00 | 0.06 |
| Optimized Z-score<sup>B</sup> |  |  |  |  |  |  |  |  |
| Anti-S1 IgG | 32.6 | 4.5 | 0.77 | 0.81 | 42.4 | 5.8 | 0.87 | 0.74 |
| Anti-NP IgG | 34.2 | 4.9 | 0.78 | 0.72 | 40.0 | 5.7 | 0.86 | 0.72 |

<sup>A</sup>Z-scores calculated for manufacturer-determined optical density ratio (ODR) positivity thresholds of 0.8 for anti-S1 IgG and 1.4 for anti-NP IgG immunoassays.
<sup>B</sup>Optimal Z-score based on the maximum value of Youden's index.

### Antibody titers over time after recovery from COVID-19

Lower nAb titers were associated with a longer time after symptom resolution according to our multivariable analysis (**Table 2**). In a cross-sectional visualization of serologic data from persons with a nAb titer of 1:40 or greater at the initial blood draw, nAb titers, anti-S1 IgG, and anti-NP IgG levels all demonstrated trends towards lower antibody levels with increased interval between symptoms and blood sampling (**Supplemental Figure 3**). To further investigate this initial observation, 41 participants enrolled during April 2020 who had nAb titers of 1:40 or greater returned for a second blood draw to examine antibody kinetics. Neutralization titers decreased over time in all but four participants. From these data, nAb half-life was calculated at 66.2 days (95% CI 55.5-82.5) (**Figure 3A**). A decline in anti-S1 IgG and anti-NP IgG levels over time was also observed in 83% and 98% of participants, respectively (**Figures 3B and 3C**). Given our observation that compression of ODRs and resultant z-scores occur at higher levels of IgG, half-life calculations were not performed for these latter assays.

**Figure 3.**
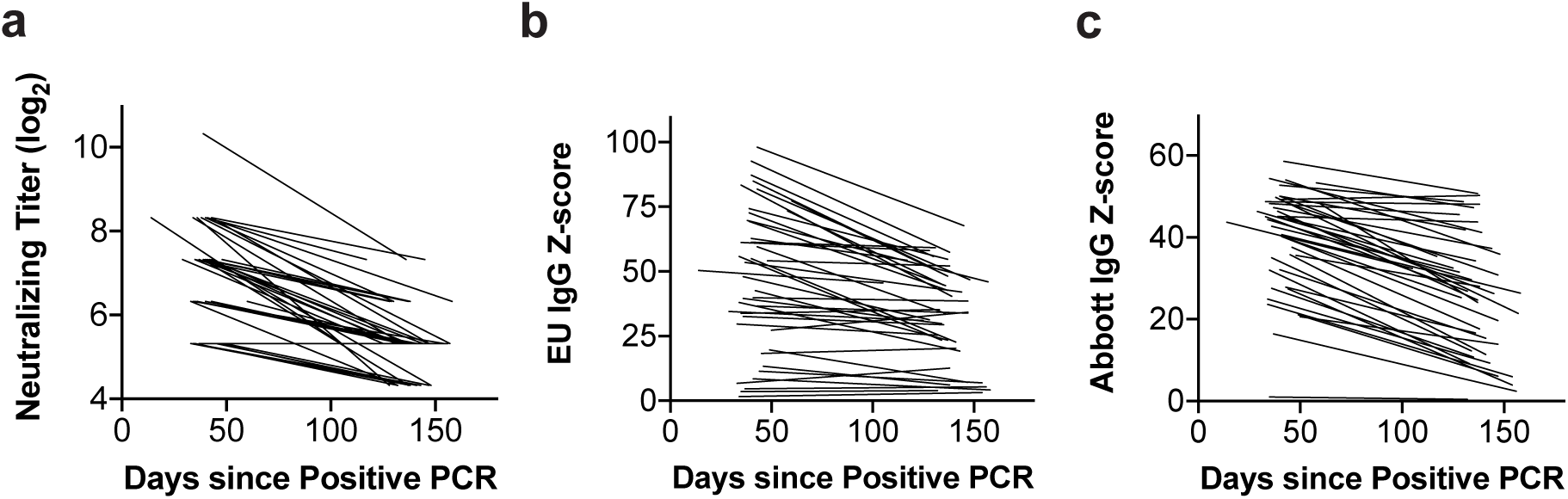
Individual antibody levels obtained at various timepoints after symptom resolution from 41 persons recovered from PCR-positive SARS-CoV-2 infection and an initial nAb titer of 1:40 or greater. The same 41 persons are represented in each graph. (A) Neutralization assay, (B) Euroimmun (anti-S1) IgG assay, (C) Abbott (anti-NP) IgG assay results as a function of time since resolution of symptoms. Each line connects two results from a single participant.

### Clinical characteristics and repeat serologic results from seronegative participants

We identified seven participants who lacked circulating antibody responses by all three assays, despite having documented SARS-CoV-2 infection. Amongst these, one had been asymptomatic and one had been hospitalized for COVID-19 (**Table 4**). Five (71%) were women with a median age of 48 (range 40-62). The median time between positive SARS-CoV-2 PCR and antibody testing was 42 days (range 30-87). The previously hospitalized participant with consistently negative antibody results across all three antibody tests had received infliximab for vasculitis one week prior to becoming infected with H1N1 influenza, followed one week later by infection with SARS-CoV-2. Six seronegative participants had a repeat blood draw, a median time of 43 days (range 20-55) after the first. Serologic test results remained negative in all individuals by all three assays (**Table 4**).

**Table 4.** Clinical and laboratory characteristics of seronegative individuals.

| Subject | Age at draw<sup>A</sup> | Sex | Severity | Comorbidities | Days sx onset to 1st draw | Days (+) PCR to 1st draw | Days symptom resolved to 1<sup>st</sup> draw | Days between 1st and 2nd draws |
| --- | --- | --- | --- | --- | --- | --- | --- | --- |
| 1 | 40s | M | Outpt | None | 41 | 38 | 30 | 54 |
| 2 | 40s | M | Outpt | None | 43 | 39 | 28 | 55 |
| 3 | 60s | F | Outpt | None | 47 | 41 | 37 | 46 |
| 4 | 40s | F | Outpt | None | 42 | 42 | 22 | 41 |
| 5 | 40s<sup>B</sup> | F | Outpt | Asthma | 52 | 50 | 40 | na |
| 6 | 40s | F | Inpt | Lung carcinoid;<br>Asthma; Vasculitis | 95 | 87 | 70 | 20 |
| 7 | 50s | F | Outpt | None | 78 | 76 | 64 | 21 |

Abbreviations: Outpt for outpatient; Inpt for inpatient; Asx for asymptomatic; sx for symptoms;
na for non-applicable
<sup>A</sup>Ages are given as decade of life
<sup>B</sup>Subject did not return for a second blood draw

To further investigate the immune status of the seronegative individuals, we performed IFN-γ ELISPOT using PBMCs from follow-up samples of the six persons with repeat blood draws, and peptides from SARS-CoV-2 structural proteins. Previous work has shown that the majority of T cell responses in COVID-19 survivors recognize these viral proteins(5). As comparators, we tested a subset of seropositive persons and pre-2019 healthy donors. We detected robust T cell responses in convalescent PBMCs from convalescent SARS-CoV-2-seropositive individuals, whereas responses from SARS-CoV-2-seronegative patients were similar to healthy donors (**Supplemental Figure 4**).

## DISCUSSION

Our characterization of a large cohort of potential SARS-CoV-2 convalescent plasma donors provides several useful observations relevant to protection after natural infection or vaccination and the manufacture of antibody-based therapeutics. We found that a significant proportion (40%) of persons failed to meet a protocol-driven nAb threshold (≥ 1:80) for contribution to a pooled immunoglobulin product. Clinical factors associated with higher levels of nAb, specifically male sex, older age, higher disease severity, and shorter interval from recovery, were identified; this information could streamline future donor recruitment. Repeat testing demonstrated a decline of functional nAb over time, suggesting that these clinical characteristics are most valuable when used early after recovery. We also found that anti-SARS-CoV-2 IgG results from two widely accessible and commercially available immunoassays could reliably predict high nAb titers when used relatively early after COVID-19. Test cutoffs were empirically derived that can assist in selecting donors with increased likelihood of having high nAb titers. Finally, approximately 3% of our cohort, all with documented SARS-CoV-2 infection, did not exhibit adaptive immune responses in multiple antibody and T cell assays.

Pending full approval of antiviral drugs or a preventative vaccine, much effort has focused on passive immunization for the treatment or prevention of COVID-19, ranging from single-donor plasma and pooled immunoglobulin products to recombinant antibodies(6-8). Pooled products have a track record of safety and efficacy for other pathogens including respiratory viruses(1), and are most efficiently manufactured from plasma containing higher amounts of functional antibody. While the immunoglobulin isotypes and functional activities associated with improved COVID-19 outcomes remain an active area of investigation(9), neutralization of viral entry is thought to be a desirable attribute that corresponds to protection. Thus, qualifying plasma donation for NIH hyperimmune globulin protocol NCT04344977 has focused on high nAb titers. Observational data from a single donor plasma program generally support the premise that neutralizing antibodies are desirable, with lower mortality rates noted for persons receiving plasma containing the top quintile of anti-S binding antibodies(6). A small randomized controlled trial also found a trend towards a shorter time to clinical improvement and lower mortality with convalescent plasma(10). We therefore sought to identify clinical characteristics associated with plasma neutralizing antibodies and compared two widely used SARS-CoV-2 antibody tests for ability to predict nAb titers of 1:80 or greater.

Our clinical findings of an association between higher nAb titers with male sex, older age, and hospitalization are consistent with a recent report from another convalescent plasma candidate donor study(11) and other reports concerning recovered individuals(12-14). The relationship between greater infection severity, and presumably greater antigen load, with higher acquired B cell responses is paralleled by data by others showing an association between more severe infection with higher T cell responses in general, and, in particular, circulating CD4+ T follicular helper (cT_FH_) responses(15). T_FH_ cells provide critical positive signals to antigen-specific B cells in lymph node germinal center (GC) reactions promoting avid and long-lasting antibody responses.

Our cross-sectional analyses also found a trend towards lower nAb titers with increased elapsed time between antibody testing after symptom resolution. This was confirmed in longitudinal analyses of 41participants re-evaluated approximately 3 months after their first test. A quarter of the re-evaluated participants had nAb titers below 1:40 at the second time point, and declines in circulating anti-S1 and anti-NP IgG levels were found in >80% of participants over the relatively short period of time between the two blood draws. A critical and currently unresolved issue for SARS-CoV-2 infection is the extent to which prior infection prevents or ameliorates various characteristics of re-infection, including disease, viral shedding, and transmission potential. It is likely that protection efficiency will vary according to clinical characteristics of the host and of the initial infection, with time after initial infection, and with the virologic or clinical endpoint under consideration. The issue of antibody durability has been addressed in prior cross-sectional and longitudinal studies, obviously limited by the recency of the pandemic. Decreases in SARS-CoV-2 nAb titers have been observed in some prior studies(13, 16, 17) but not others(18). The decline of nAb titers over time and the low somatic hypermutation in immunoglobulin genes recovered from SARS-CoV-2-specific memory B cells(19) both suggest that ineffective help from T_FH_ CD4 T cells is occurring in germinal centers. While circulating T_FH_-like SARS-CoV-2-reactive CD4 T cells have been detected in the blood(15), autopsy data from fatal COVID-19 is consistent with poor germinal center reactions(20). Overall, waning immunity after natural infection raises concern for re-infection over time as observed for other respiratory viruses, including the endemic coronaviruses(2, 3). Data on protection from re-infection with SARS-CoV-2 are incomplete at this time, with case reports of re-infection(21) contrasting with cohort data from an occupational outbreak in which persons with baseline SARS-CoV-2-specific circulating antibody were protected from infection(22). Additional research will be required to address the immunologic and clinical parameters associated with protection from re-infection after SARS-CoV-2 infection.

With regard to test performance, we found a slightly stronger correlation between nAb titers and anti-S1 IgG levels using the Euroimmun assay compared to anti-NP IgG levels determined using the Abbott assay. We were able to establish cutoffs using z-scores for each assay that had good performance for predicting nAb titers of 1:80 or greater, or 1:160 or greater, which should be useful as the clinical efficacy of single-donor and pooled immunoglobulin products continue to be defined. The superior performance of the anti-S1 assay is not unexpected given that antibodies that neutralize SARS-CoV-2 bind to the S protein present on the surface of infectious virus. This finding is also consistent with studies that have shown that the detection of NP-binding antibodies does not always correlate with the presence of nAb(23).

In the present study, we used z-scores to standardize serologic results to the same scale and to account for local test performance characteristics(24). Despite using a relatively conservative positivity threshold of 3 standard deviations above our assay-specific negative control means, these z-score positivity thresholds were lower than manufacturer-recommended thresholds for both assays. Accordingly, a larger number of samples were considered “negative” by both assays using manufacturer-recommended thresholds compared to z-scores. Although continuous values are obtained and accessible for both Abbott and EU assays, it is worth noting that both are EUA-approved as qualitative and not quantitative assays. Although the Euroimmun assay appears to be have a wider analytic range in comparison to the Abbott assay, both assays yield results that plateau at different thresholds. Thus, values at higher levels for both assays would require dilution of high-titer sera to achieve linear results. However, even in the absence of re-assay after dilution, however, we were able to discern declines in binding IgG titers over time within-subjects, similar to our observations for nAb.

A small proportion (3%) of participants tested negative on all three antibody assays despite SARS-CoV-2 RNA detection in previously collected respiratory specimens. Each seronegative COVID-19 survivor tested also had low T cell responses, similar to pre-2019 healthy controls, while seropositive subjects had abundant interferon-gamma producing cells in the blood, similar to other reports(25). While false positive PCR results are possible, all subjects except one, who was a household contact of a person with symptomatic and PCR-confirmed COVID-19, had a clinical syndrome consistent with COVID-19. Asymptomatic and mildly symptomatic patients with COVID-19 have been found to have lower antibody levels and also were more likely to sero-revert in the early convalescent phase(14). Discordance between positive PCR and negative circulating antibody may be related to timing of the blood draw after illness, milder symptoms correlating with lower or briefer antigen exposure, and/or an unrecognized immune suppression.

Our study has some limitations. First, although our sample population included a clinically diverse population with respect to age, sex, and clinical presentation, the cohort was predominantly white. Additional studies in minority populations at risk for worse outcomes will be needed to determine the generalizability of these results. Second, our study was mainly cross-sectional, and longitudinal data were obtained on only a subset of the cohort. Third, in analyzing the decline of neutralizing antibody titers, a titer of 1:20 was used as a surrogate for all titers <1:40, since this value was the next serial 2-fold titer below the lowest positive result. We chose this conservative surrogate value to avoid the possibility of over-estimating the rate of decline in neutralizing antibody titers. Fourth, we focused on blood immune responses. It is possible that the seronegative individuals in our study mounted a mucosal humoral or T cell response that could provide protection against re-infection. Probing mucosal and T cell immunity may be important in understanding the immune response of subjects with milder infection.

In summary, our data provide important information regarding the predictive value of clinical factors and commercially available immunoassay results for high nAb titers. This information could aid in streamlining the selection of potential convalescent plasma donors and increase access to and efficiency of donor sample testing. Since transfusion of plasma with higher antibody titers has been associated with improved outcomes(6), and since higher titer plasma is the most suitable for the creation of pooled products, our data further suggest that plasma donations should be sought closer to the resolution of symptoms, rather than later, to optimize yield. A minimum level of nAb necessary for protection has not been established and it is unknown whether persons with low titers will boost their immune responses upon re-exposure. However, our data raise concern that functional antibodies show a quantitative decline over a relatively short period of time after recovery in those with relatively mild infections. Further insight into these issues is critical to monitoring herd immunity and implementation of immunization programs.

## METHODS

### Study description

Seattle-area participants were recruited for potential donation of single donor plasma units (NCT04338360), and plasma for manufacture of a pooled anti-SARS-CoV-2 product (NCT04344977). Both studies required age 18 or greater and PCR-positivity to SARS-CoV-2; criteria for donation for the pooled product also included a nAb titer of ≥ 1:80(26). Participants provided demographic and clinical information, including positive SARS-CoV-2 results, and a blood sample.

### Serologic assays

Plasma from EDTA and heparinized blood and serum were prepared within 12 hours of collection. Serum and plasma for IgG assays were stored at 4°C and assays performed within 72 hours of sample processing. PBMCs were isolated from heparinized blood within 12 hours of phlebotomy using standard Ficoll-Hypaque density gradient centrifugation and cryopreserved in liquid nitrogen in 90% fetal calf serum/10% DMSO. EDTA plasma was stored at −80°C for neutralization assays.

Serologic testing was performed using two FDA-authorized (via Emergency Use Authorization), CE-marked immunoassays; both are approved to provide qualitative results, although both yield quantitative values. The anti-SARS-CoV-2 IgG ELISA from Euroimmun (Lubeck, Germany) measures antibodies to recombinant spike protein (S1 domain), containing the receptor binding domain that interacts with ACE2(27). All testing and analyses were performed according to the manufacturer’s protocols, with the OD ratio (ODR) calculated using the kit calibrator. The manufacturer-provided reference range is as follows: ratio <0.8 (negative), ratio ≥ 0.8 to <1.1 (borderline), and ratio ≥ 1.1 (positive). The Abbott SARS-CoV-2 IgG chemiluminescent microparticle immunoassay (Abbott ARCHITECT, Abbott Park, IL, USA) measuring antibodies to the SARS-CoV-2 nucleoprotein (NP) was performed per the manufacturer’s instructions. Equivalent to an ODR, the kit calibrator was used to establish the assay index result. Index values ≥ 1.4 are considered positive by the manufacturer.

To standardize results and facilitate comparisons, ODR scores for each sample were converted to z-scores (number of SDs above the negative control mean) as follows: z-score = (Test ODR – Mean negative control ODR) / Mean negative control standard deviation (SD)(24). Negative control sera had been collected between 2015 and 2019 from healthy community blood donors and from patients tested in the clinical laboratory by western blot for potential HSV infection (N=78 for Euroimmun and N=136 for Abbott). ODR were converted to z-scores as follows: For Euroimmun, z-score = (ODR – 0.26)/0.13 and for Abbott, z-score = (index result – 0.08)/0.14). A conservative z-score ≥ 3 was considered positive to minimize false positive results.

Neutralizing antibody titers were performed using a fluorescence reduction neutralization assay (FRNA) with authentic SARS-CoV-2 (2019-nCoV/USA-WA1-A12/2020 from the US Centers for Disease Control and Prevention, Atlanta, GA, USA) at the NIH-NIAID Integrated Research Facility at Fort Detrick, MD, USA. In these experiments, a fixed volume of diluted virus was incubated with an equivalent volume of test plasma for 1 hour at 37 °C prior to adding the suspension to Vero E6 cells (ATCC, Manassas, VA, USA, #CRL-1586). The virus was allowed to propagate for 24 hours prior to fixing the cells. Following fixation, the cells were permeabilized and probed with a SARS-CoV-2 nucleoprotein-specific rabbit primary antibody (Sino Biological, Wayne, PA, USA, #40143-R001) followed by an Alexa594-conjugated secondary antibody (Life Technologies, San Diego, CA, USA, #A11037). The total number of infected cells in four fields per well with each field containing at least 1000 cells was quantified using an Operetta high content imaging system (Perkin Elmer, Waltham, MA, USA). Plasma was tested using two-fold serial dilutions from 1:40 to 1:1280 with four replicates per dilution. Results are reported as the highest dilution of plasma leading to at least 50% reduction of SARS-CoV-2 titers. Each assay was controlled with internal addition of an S-specific neutralizing polyclonal antibodies. If a 1:40 dilution did not lead to at least 50% reduction of viral titer, results were reported as <1:40 even though some inhibition of virus propagation may have been present. To enable analysis of nAb kinetics over time, a value of 1:20 was used as a surrogate for results <1:40 since this value was the next serial 2-fold titer below the lowest positive result.

### T cell assays

To measure T cell responses to SARS-CoV-2, PBMCs were thawed and plated at 2.5-3 × 10^5^ cells/well in duplicate. Stimuli were 9 pools of SARS-CoV-2 peptides (Genscript, Piscataway, NJ, USA) corresponding to the S, membrane (M), and NP proteins of SARS-CoV-2 strain Wuhan (Genbank MN908947.3) in pools of 50-56 peptides/pool. Peptides 13 amino acids long and overlapping by 9 amino acids were used at a final concentration of 1 μg/mL for each peptide and 0.2% DMSO. Negative control was 0.2% DMSO; positive control was 1.6 μg/mL phytohemagglutinin (PHA-P, Remel, Lenexa, KS, USA). Interferon-gamma (IFN-γ) ELISPOT was performed as described(28). All subjects appropriately had too-numerous-to-count positive responses to PHA. Results are reported as spot-forming units/10^6^ PBMCs for the background-subtracted response to SARS-CoV-2 peptide pools.

### Statistics

Univariable analysis was performed using a t-test for continuous variables and Chi-square test for categorical variables. Variables with *P*≤0.2 in univariable analysis were candidates for multivariable models using logistic regression and were retained in the models if they remained significant or substantially modified the effect of another factor. Statistical significance was defined as two-sided *P*<0.05. The sensitivity and specificity for high nAb titers by the Abbott and the Euroimmun assays were plotted by generating a receiver operating characteristic (ROC) curve. SPSS software (IBM, Chicago, IL, USA) was used for statistical analysis and to generate ROC curves, the latter with default parameters. Youden’s index was calculated for all points of the ROC curves, and the maximum index value was used to select the optimal z-score cut-off(4). Longitudinal analysis of nAb levels was performed using linear mixed effects models with the R package *lme4*(29).

### Study approval

Institutional Review Board approval was obtained from the University of Washington and all participants underwent informed consent form.

## Data Availability

Relevant data are available from the corresponding author on request.

## ABBREVIATIONS

DMSO: dimethylsulfoxide
PCR: polymerase chain reaction
EDTA: ethylenediamine tetra-acetic acid
SARS-CoV-2: severe adult respiratory syndrome coronavirus 2
COVID-19: coronavirus disease 2019
OR: odds ratio
CI: confidence interval
IgG: immunoglobulin G
BSL-3: biosafety level 3
nAb: neutralizing antibody
ROC: receiver operating curve
S1: spike protein domain 1
NP: nucleoprotein
PBMC: peripheral blood mononuclear cells
ACE2: angiotensin converting enzyme 2
CE: Conformité Européenne
ELISA: enzyme linked immune sorbent assay
ODR: optical density ratio
FRNA: fluorescence reduction neutralization assay

## AUTHOR CONTRIBUTIONS

J.B., C.M., M.K.D., D.M.K, and A.W. wrote the manuscript with input from all co-authors. A.W. and T.B.G. conceived the study. J.B. and A.W. wrote the IRB protocol. J.B., D. Z., S. McGuffin, and A.E.S., recruited participants. S.S. managed the data. V.L.C., C.L.M., and D.M.K. processed specimens. C.M., A.C., M.K.D., A.G., K.R.J., S.L.F., L.J., C.L.M., and M.W. characterized and performed antibody and ELISPOT immunoassays. M.H., R.G., J.L., E.P., and S.M. performed the neutralization assays. J.B., C.M., S.S., M.W., A.W., and D.M.K. performed statistical analysis and generated tables and figures. The order of the first author is alphabetical.

## ACKNOWLEDGMENTS

We are grateful to all study participants. We thank Angela LeClair, Miko Robertson, Mark Drummond, Kirsten Hauge, Isabelle Hwang, Kristina Chaffee, Avery Forrow, and Tanvi Aggarwal for support in recruiting and enrolling subjects. We thank the members of the University of Washington Clinical Immunology and Virology Labs for conducting the serological testing.

