## Supplemental Figures for "Clinical, laboratory, and temporal predictors of neutralizing antibodies to SARS-CoV-2 after COVID-19"

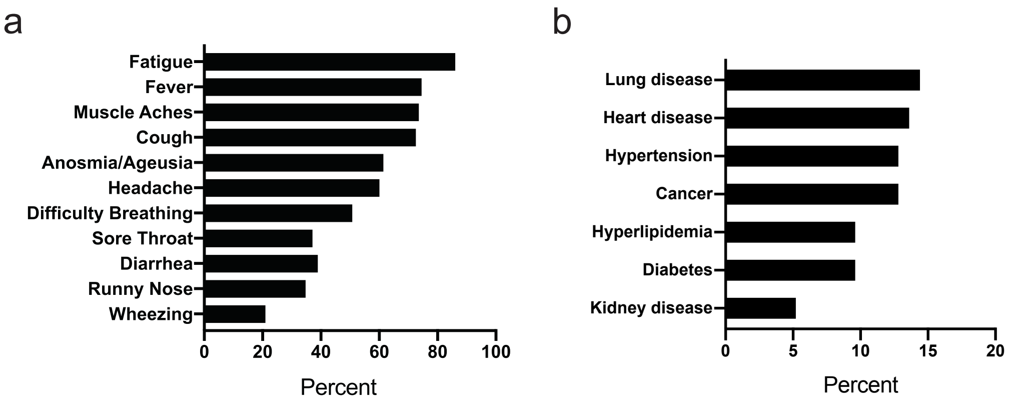


**Supplemental Figure 1.** Prevalence of symptoms (A) and comorbidities (B) in SARS-CoV-2-infected persons screened for convalescent plasma donation.

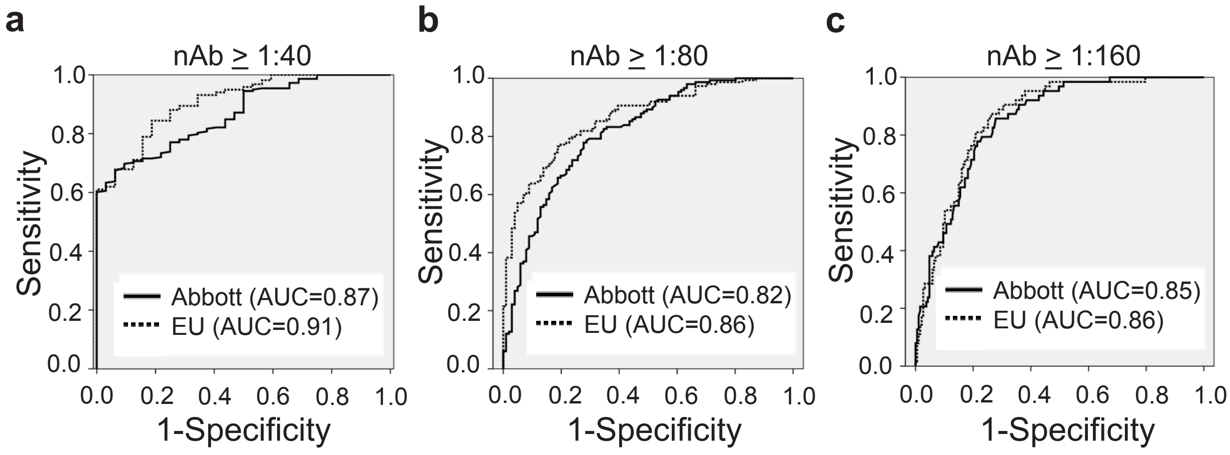


**Supplemental Figure 2.** Receiver operating characteristic (ROC) curves for predicting nAb levels. ROC curves for nAb titers (A) ≥ 1:40, (B) ≥ 1:80, and (C) ≥ 1:160 based on z-scores from the Abbott and Euroimmun assays. Abbreviations: nAb, neutralizing antibody; AUC, area under the curve; EU, Euroimmun.

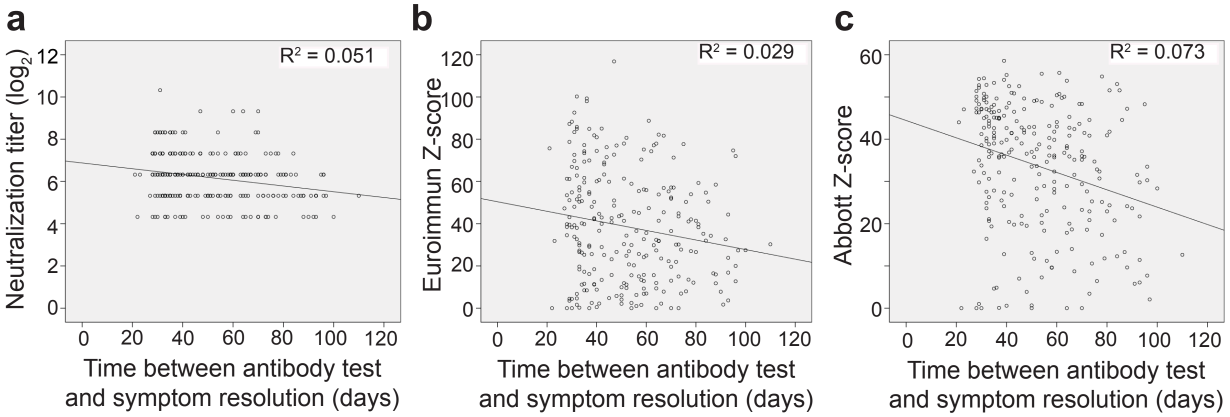


**Supplemental Figure 3.** Cross-sectional analyses of candidate COVID-19 convalescent plasma donors with nAb titers of 1:40 or greater at the time of initial blood draw. The X axis is the number of days between the resolution of COVID-19 symptoms and the time of blood sampling. The dotted line represents the best fit by linear regression.

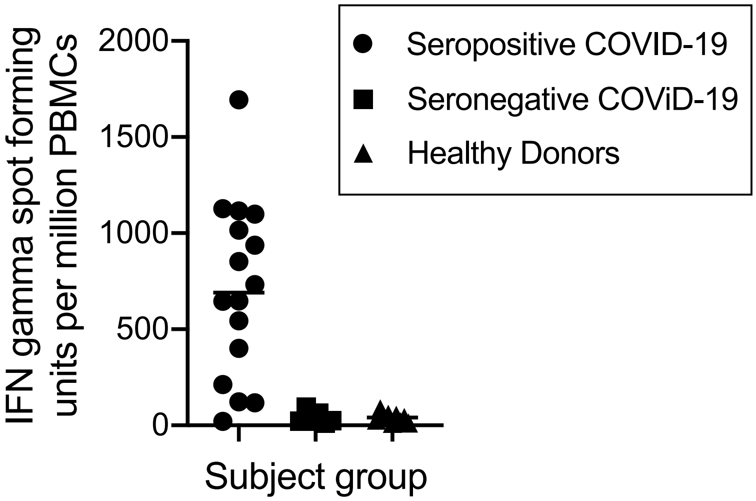


**Supplemental Figure 4.** PBMC interferon gamma ELISPOT responses to SARS-CoV-2 peptides covering the major structural NP, S, and membrane proteins. Each symbol represents a single donor with the serologic and clinical characteristics indicated.
